## Supplemental Information for "Socioeconomic context influences the heritability of child cortical structure"

**SI Results**

*Genetic contribution to lobar cortical structure*

Lobar analyses revealed that the heritability of cortical thickness ranged from 0.69 to 0.80, surface area from 0.69 to 0.90, sulcal depth from 0.34 to 0.61, and GWC from 0.48 to 0.59 (SI Table 1).

| Imaging metric and lobe | Heritability estimate |
| --- | --- |
| Thickness cingulate | 0.69 |
| Thickness frontal | 0.80 |
| Thickness occipital | 0.74 |
| Thickness parietal | 0.77 |
| Thickness temporal | 0.74 |
| Area cingulate | 0.69 |
| Area frontal | 0.90 |
| Area occipital | 0.75 |
| Area parietal | 0.74 |
| Area temporal | 0.84 |
| Sulcal depth cingulate | 0.41 |
| Sulcal depth frontal | 0.61 |
| Sulcal depth occipital | 0.54 |
| Sulcal depth parietal | 0.42 |
| Sulcal depth temporal | 0.34 |
| GWC cingulate | 0.54 |
| GWC frontal | 0.58 |
| GWC occipital | 0.46 |
| GWC parietal | 0.48 |
| GWC temporal | 0.59 |

SI Table 1. Lobar heritability per metric.

***Heritability of cortical structure within genetically homogeneous sample***
Overall heritability estimates of cortical structure within the more genetically homogeneous subsample were highly similar to the original analyses (SI Table 2). SES moderation analyses also produced results consistent with the main findings, apart from surface area, where the effect weakened (SI Table 3). This may either reflect reduced power in the sensitivity analysis or inflated (albeit nonsignificant) moderation in the original sample due to population stratification.

| Global imaging metric | Original sample (n = 9080) | Ancestrally homogeneous sample (n = 7445) |
| --- | --- | --- |
| Thickness | H^2^ = 0.82 | H^2^ = 0.83 |
| Area | H^2^ = 0.82 | H^2^ = 0.88 |
| Sulcal depth | H^2^ = 0.57 | H^2^ = 0.59 |
| GWC | H^2^ = 0.57 | H^2^ = 0.53 |

SI Table 2. The table shows the heritability estimates for the global imaging metrics in the original sample and in the ancestrally homogeneous subsample.

| Metric | Original sample (n = 9080) | Homogeneous sample (n = 7445) |
| --- | --- | --- |
| Thickness | χ² = 8.27, p = 0.041, pFDR = 0.051 | χ² = 10.94, p = 0.012, pFDR = 0.024 |
| Area | χ² = 8.34, p = 0.040, pFDR = 0.051 | χ² = 0.01, p = 1.000, pFDR = 1.000 |
| Sulc. depth | χ² = 1.08, p = 0.781, pFDR = 0.781 | χ² = 1.25, p = 0.740, pFDR = 0.987 |
| GWC | χ² = 13.59, p = 0.004, pFDR = 0.010 | χ² = 16.24, p = 0.001, pFDR = 0.004 |

SI Table 3. The table shows the chi-squared values and the accompanying p-values for the global imaging metrics in the original sample and in the ancestrally homogeneous subsample.

*Heritability of cortical structure using twin-based assessments*

**Heritability estimates for all global imaging metrics, calculated using Falconer’s formula applied to twin-based intra-class correlations, were generally aligned with SNP-based estimates but slightly lower overall - except for mean sulcal depth, which was somewhat higher. The twin-based estimates were: cortical thickness = 0.71, surface area = 0.77, sulcal depth = 0.67, and GWC = 0.45.**

**SI Methods and Materials**

*Participants*

All ABCD data is stored in the NIMH Data Archive Collection #2573, which is available for registered and authorized users (Request #7474, PI: Westlye). ABCD Study^®^ exclusion criteria included non-English proficiency, contraindications for MRI, a history of major neurological disorders, extremely premature birth, and a diagnosis of schizophrenia, substance abuse disorder or moderate to severe autism spectrum disorder (Karcher et al., 2018). 1934 individuals were excluded due to missing data on either initial demographic, socioeconomic, neighborhood, cognitive, genetic ethnicity, family ID, scanner, imaging quality, or cortical macro- or microstructure-based variables of interest (SI Figure 1). SI Figure 2 shows the spread of SES and cognitive abilities for excluded individuals.

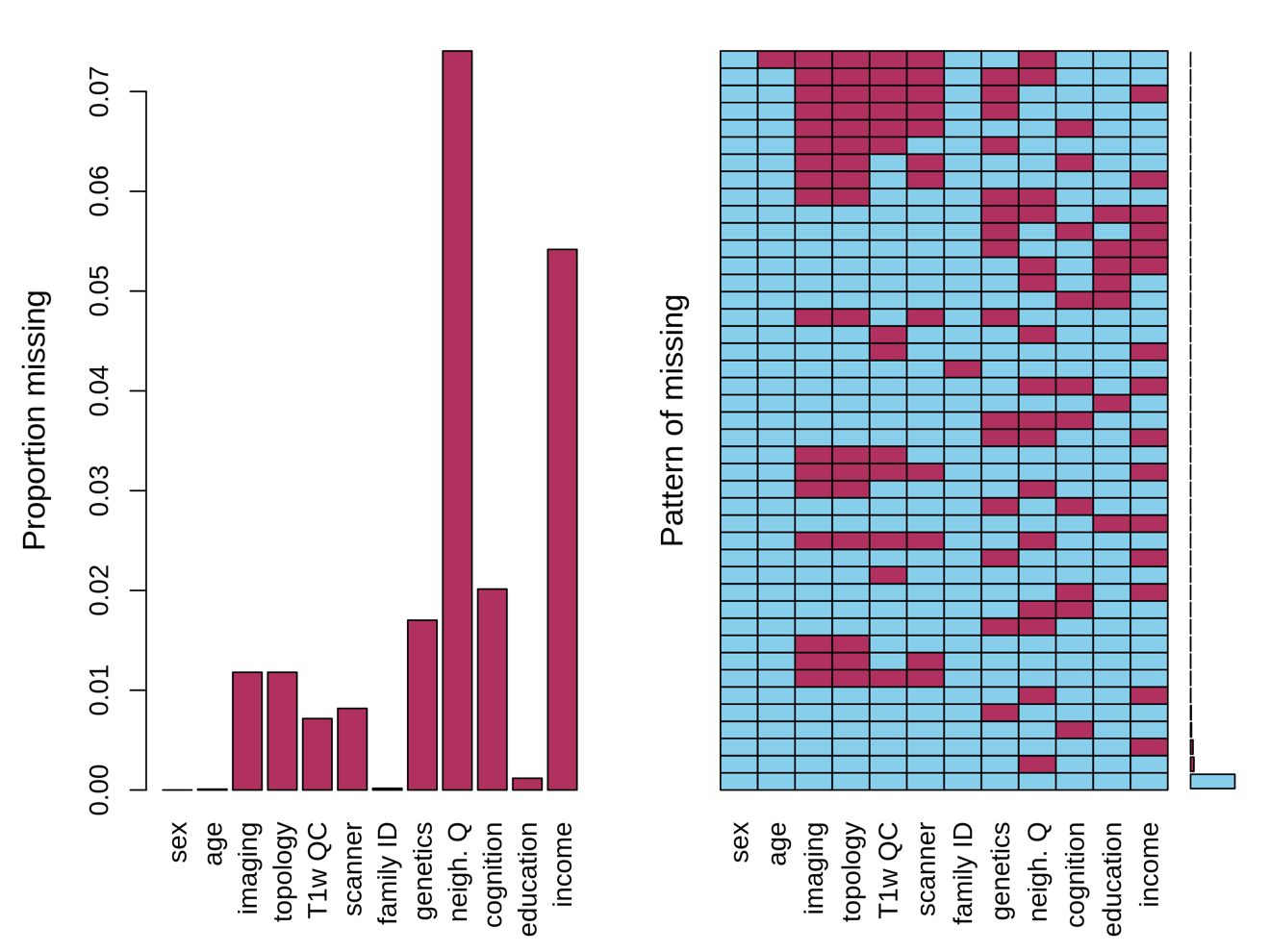

**SI Figure 1.** Proportion and pattern of missing data (NAs) across demographic, socioeconomic, neighborhood, cognitive, genetic ethnicity, family ID, scanner, imaging quality, or cortical macro- or microstructure-based variables of interest. The left panel shows the proportion of missing values for each variable. The right panel displays the missing data pattern, with blue indicating observed values and red indicating missing values.

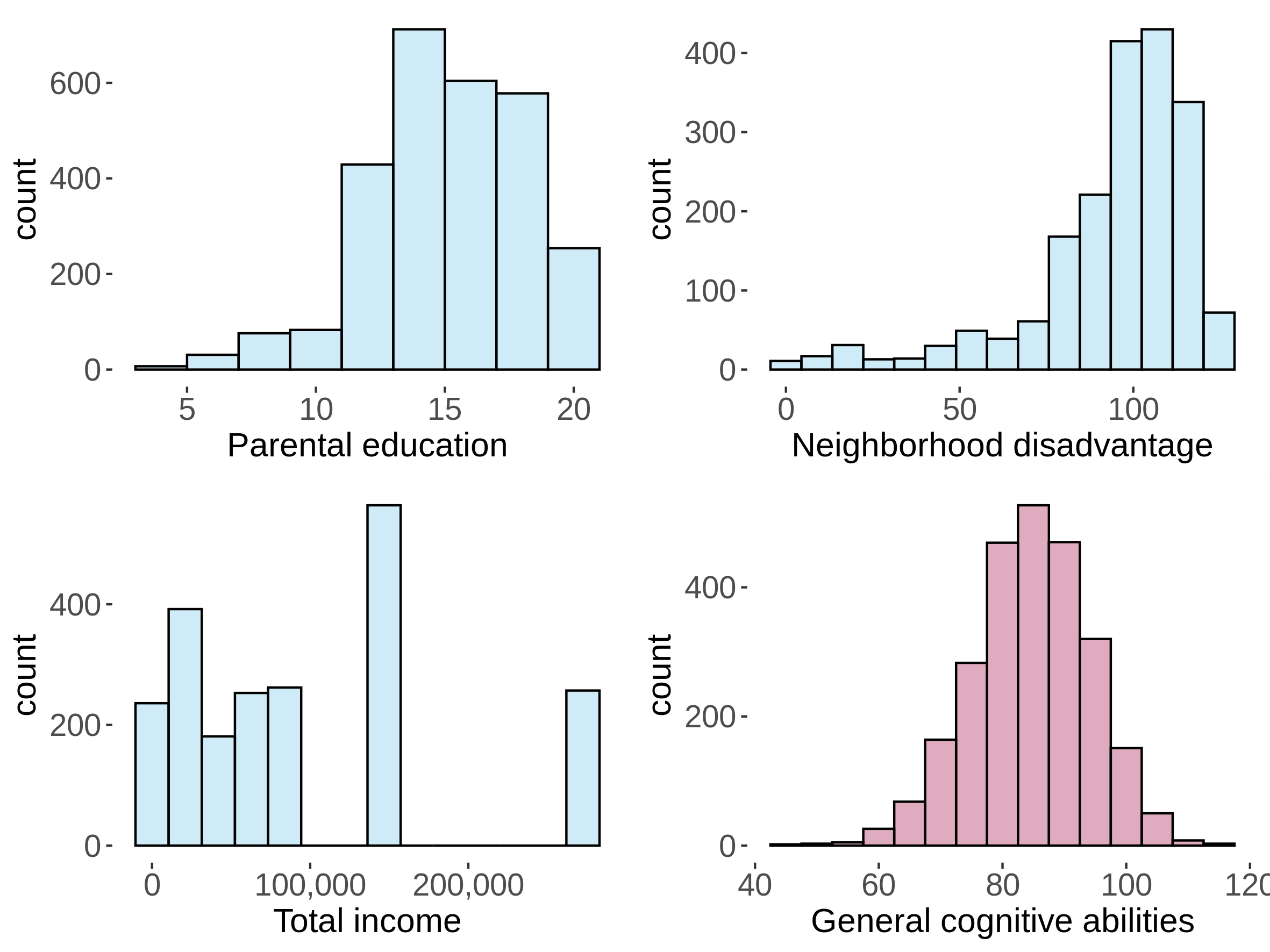

**SI Figure 2.** Distribution of socioeconomic status (SES) indicators and general cognitive abilities for all excluded individuals (n = 2788). Participants with missing values on the respective variables are not shown.

*Measurement of socioeconomic and neighbourhood deprivation*

A partner to the parent was defined as “any significant figure that helps you in raising your child or has helped you for more than 2 years and is =>40% involved in the child’s daily activities, i.e., a spouse, boyfriend/girlfriend or relative”. See SI Table 2 for parental education and income details as well as recoding.

The first component of the PCA which explained 65% of the total variance, was extracted as the overarching “SES-index” with contributions from income, education, and neighborhood deprivation in that order. Higher values reflect higher income and education, and lower neighborhood deprivation (SI Figure 3). See SI Figure 4 for the distribution of SES and neighborhood-related variables in the final sample.

Despite the availability of the “ABCD Occupation Survey Parent” questionnaire (https://nda.nih.gov/data_structure.html?short_name=abcd_occsp01), we decided to not incorporate the well-known SES subfactor “occupation”. While the questionnaire is detailed and includes occupations on a 22-point scale, it does not attempt to order them based on status. To maintain the integrity of our SES measure and avoid introducing subjective bias, we refrained from attempting to assign our own rankings to reported occupations. As an example, it is also unclear how to hierarchically rank the first 5 categories: 1=Management; 2=Business and Financial Operations; 3=Computer and Mathematics; 4=Architecture and Engineering; 5=Life, Physical and Social Sciences.

| Question | Representation of numbers | Recoding |
| --- | --- | --- |
| What is the highest grade or level of school you have completed or the highest degree you have received? | 0= Never attended/Kindergarten only  1= 1st grade  2= 2nd grade  3= 3rd grade  4= 4th grade  5= 5th grade  6= 6th grade  7= 7th grade  8= 8th grade 8  9= 9th grade  10= 10th grade  11= 11th grade  12= 12th grade  13= High school graduate  14= GED or equivalent  15= Some college  16= Associate degree: Occupational  17= Associate degree: Academic Program  18= Bachelor's degree (i.e. BA)  19= Master's degree (i.e. MA)  20= Professional School degree (i.e. MD)  21= Doctoral degree (i.e. PhD) | 0= 0 years of education  1= 1 year of education  2= 2 years of education  3= 3 years of education  4= 4 years of education  5= 5 years of education  6= 6 years of education  7= 7 years of education  8= 8 years of education  9= 9 years of education  10= 10 years of education  11= 11 years of education  12= 12 years of education  13= 12 years of education  14= 12 years of education  15= 14 years of education  16= 14 years of education  17= 14 years of education  18= 16 years of education  19= 18 years of education  20= 20 years of education  21= 21 years of education |
| How much did you earn, before taxes and other deductions, during the past 12 months? | 1= Less than $5,000  2= $5,000 through $11,999  3= $12,000 through $15,999  4= $16,000 through $24,999  5= $25,000 through $34,999  6= $35,000 through $49,999  7= $50,000 through $74,999  8= $75,000 through $99,999  9= $100,000 through $199,999;  10= $200,000 and greater  777= Refuse to answer  999= Don't know | 1= $2500  2= $8499,5  3= $13999,5  4= $20499,5  5= $29999,5  6= $42499,5  7= $62499,5  8= $87499,5  9= $149999,5  10=275000  777= NA  999= NA |
| What is your total combined family income for the past 12 months? This should include income from all sources, wages, rent from properties, social security, disability and/or veteran's benefits, unemployment benefits, help from relative and so on | 1= Less than $5,000  2= $5,000 through $11,999  3= $12,000 through $15,999  4= $16,000 through $24,999  5= $25,000 through $34,999  6= $35,000 through $49,999  7= $50,000 through $74,999  8= $75,000 through $99,999  9= $100,000 through $199,999;  10= $200,000 and greater  777= Refuse to answer  999= Don't know | 1= $2500  2= $8499,5  3= $13999,5  4= $20499,5  5= $29999,5  6= $42499,5  7= $62499,5  8= $87499,5  9= $149999,5  10=275000  777= NA  999= NA |

SI Table 4. Parental education and parental income details. The table shows the socioeconomic status related questions presented to the parents, its numeric levels, and what these levels represent.

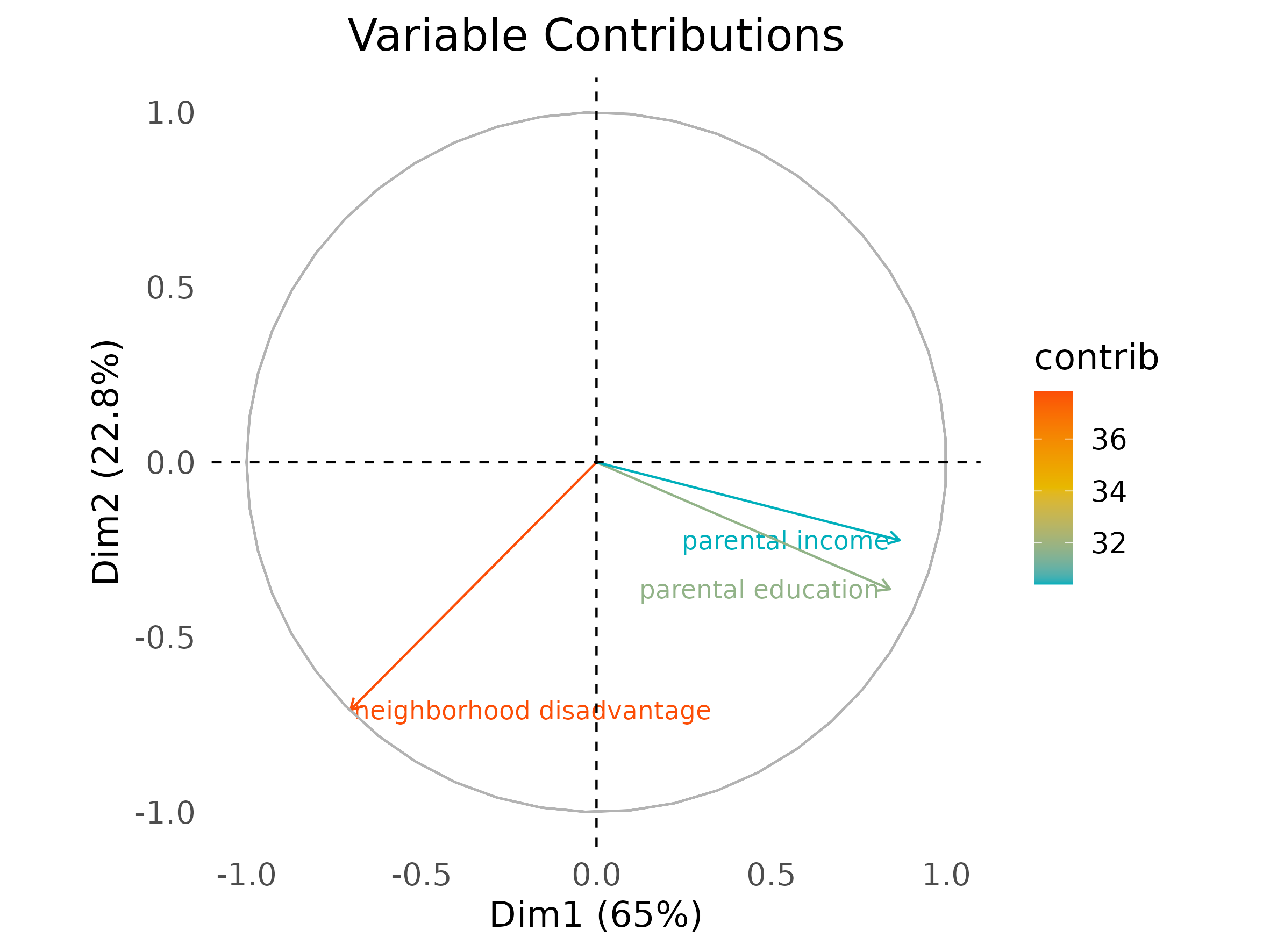

**SI Figure 3.** Biplot showing the contribution and orientation of socioeconomic status (SES) subfactors. The plot displays parental income, parental education, and neighborhood disadvantage across dimensions 1 and 2, representing the first two principal components.

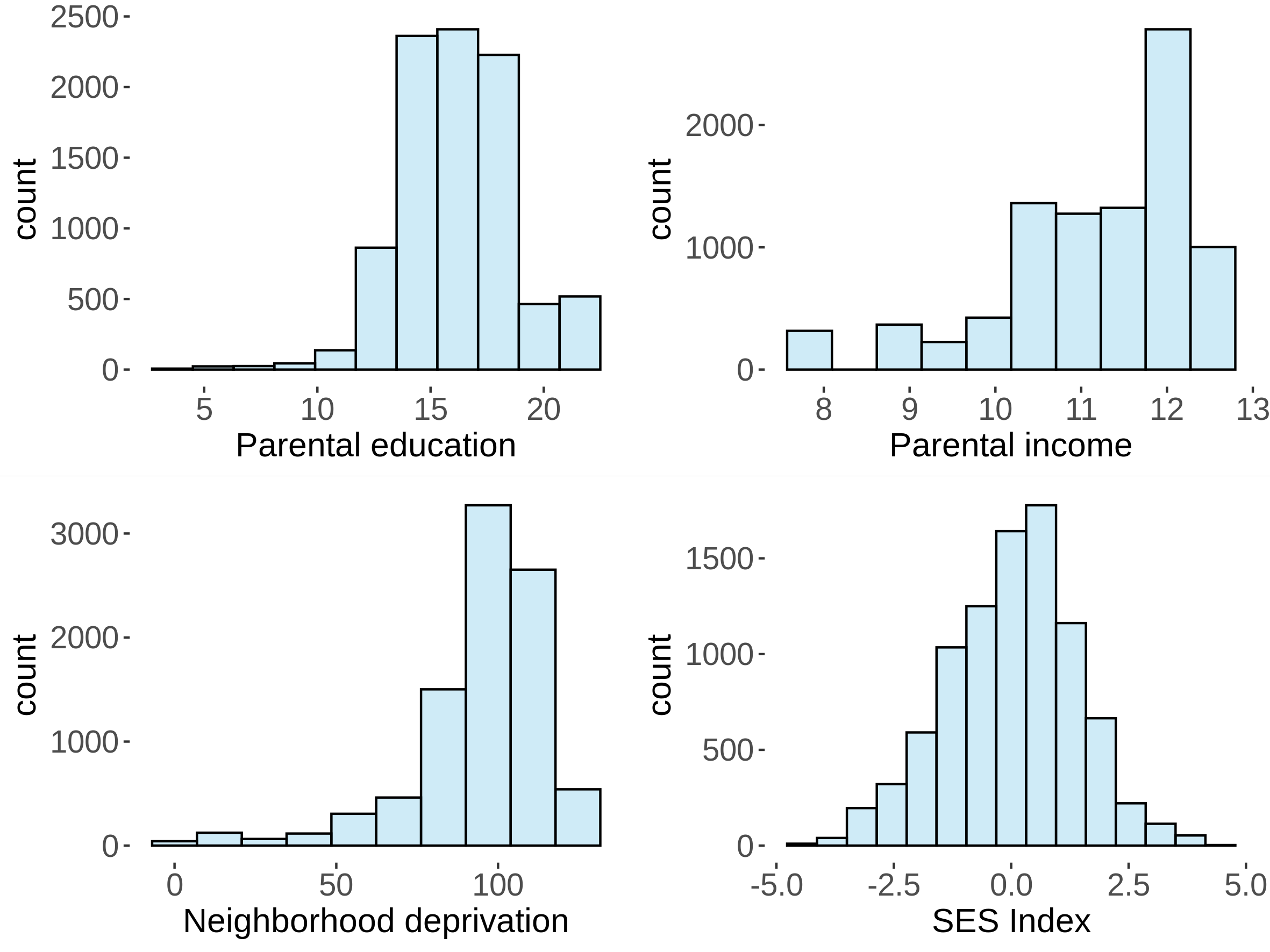

SI Figure 4. Distribution of socioeconomic status (SES) subfactors and the SES-index within the final sample. Histograms show the spread of parental education, parental income, neighborhood deprivation, and the derived SES index within the final sample (n= 9080).

*Construction of a genetically homogeneous subsample*

To mitigate residual population stratification, we constructed a more genetically homogeneous subsample using the genetic principal components (PCs). We examined the first 10 PCs and retained those whose correlation with the SES index was at least 0.10 in magnitude (PCs 1–5 and 7; SI Figure 5). We then applied k-means clustering (k=2; nstart=25) and retained the larger cluster as the genetically homogeneous subsample (n=7,445). The excluded cluster appeared to be disproportionately of lower-SES (Blue group; SI Figure 6) and predominantly parent-reported as Black (1,189). In contrast, the included subgroup was parent-reported predominantly as White (5,063) and Hispanic (1,597), contained all Asian participants (120), and had only 16 Black participants (SI Table 5).

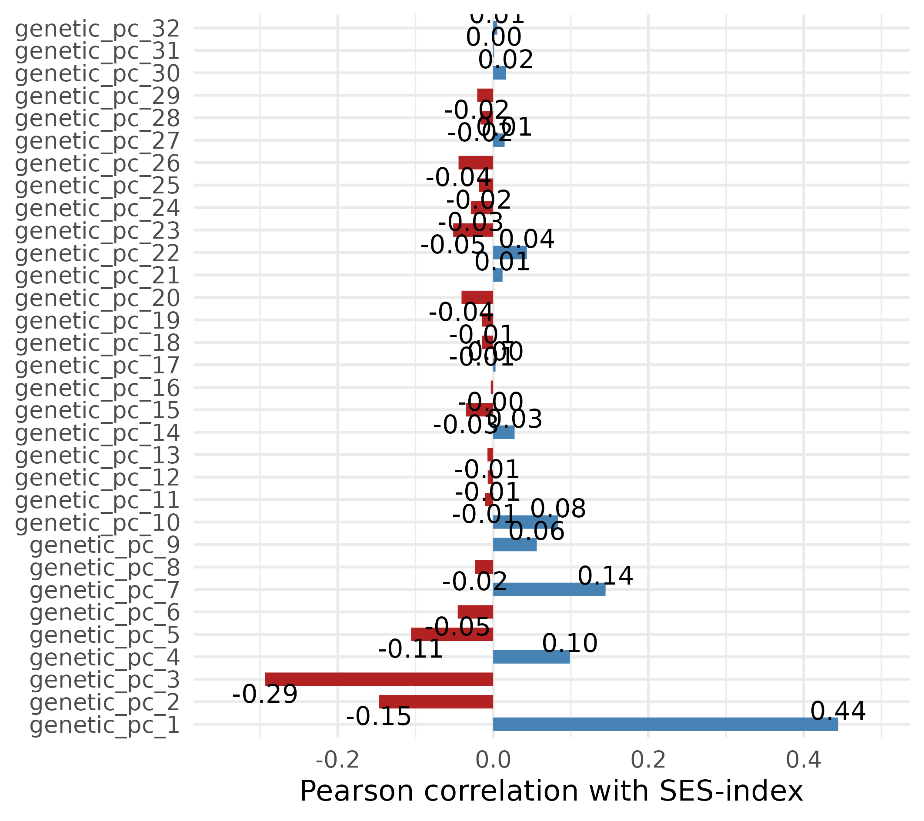

SI Figure 5. Pearsons’s correlations of each of the 32 genetic principal component and the SES-index.

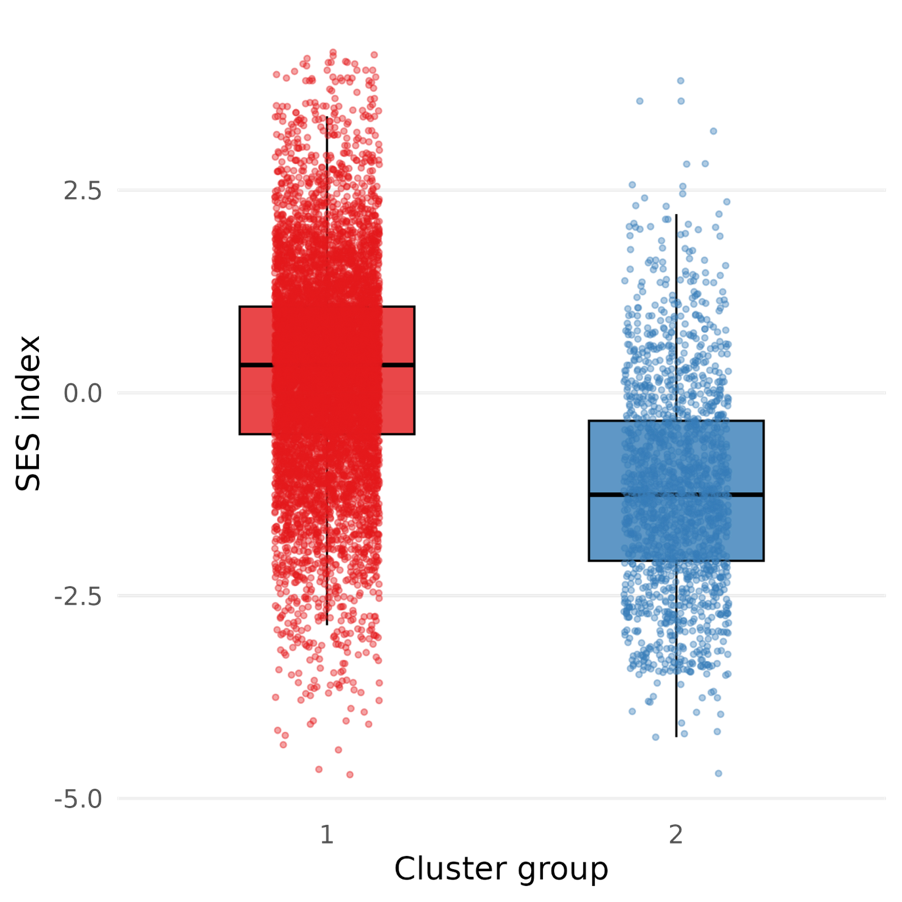

SI Figure 6. Box plot of the distribution of the SES-index across the homogeneous subsample (group 1) and the excluded individuals (group 2).

| Cluster | Asian | Black | Hispanic | Other | White |
| --- | --- | --- | --- | --- | --- |
| 1 | 120 | 16 | 1597 | 649 | 5063 |
| 2 | 0 | 1189 | 120 | 323 | 3 |

SI Table 5. Parent-reported ethnicity for group 1, included in the sensitivity analyses as a more genetically homogeneous subgroup, and for group 2, which was excluded from these analyses.

*Twin-based assessments of heritability*

As a confirmatory analysis, we additionally estimated heritability using Falconer’s formula based on intraclass correlations in monozygotic (MZ, n = 506) and dizygotic (DZ, n = 788) twins. Zygosity was determined using pedigree-based familial relationships, and only complete twin pairs were included in the analyses.

*Scanner harmonization*

To adjust for systematic and unwanted scanner-related variance, imaging metrics were subsequently imported to R and the package neuroCombat (Fortin et al., 2018) was employed to harmonize data across each of the 29 scanners. We included relevant covariates to our model, namely age, sex, parental education, parental income, neighborhood deprivation, general cognitive ability, family ID, and 32 genetic principal components (PCs), to preserve such variance during the harmonization procedure. Box plots of global MRI measures pre- and post neuroCombat adjustments are presented in SI Figure 7-10.

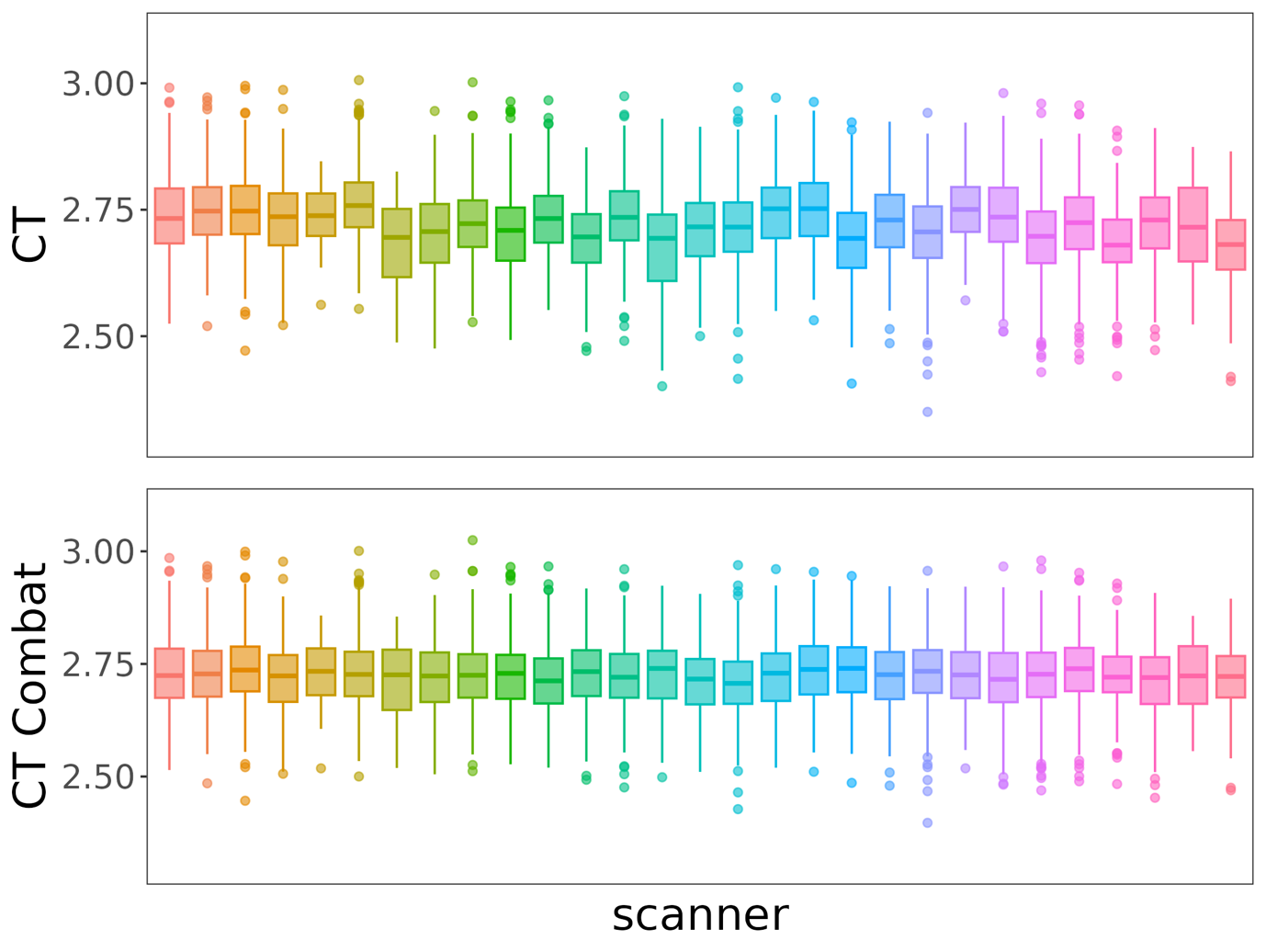
SI Figure 7. Box plots of mean cortical thickness across scanners pre- and post harmonization. The figure shows box plots of the distribution of mean cortical thickness across scanners pre-scanner harmonization at the top, and post-harmonization at the bottom.

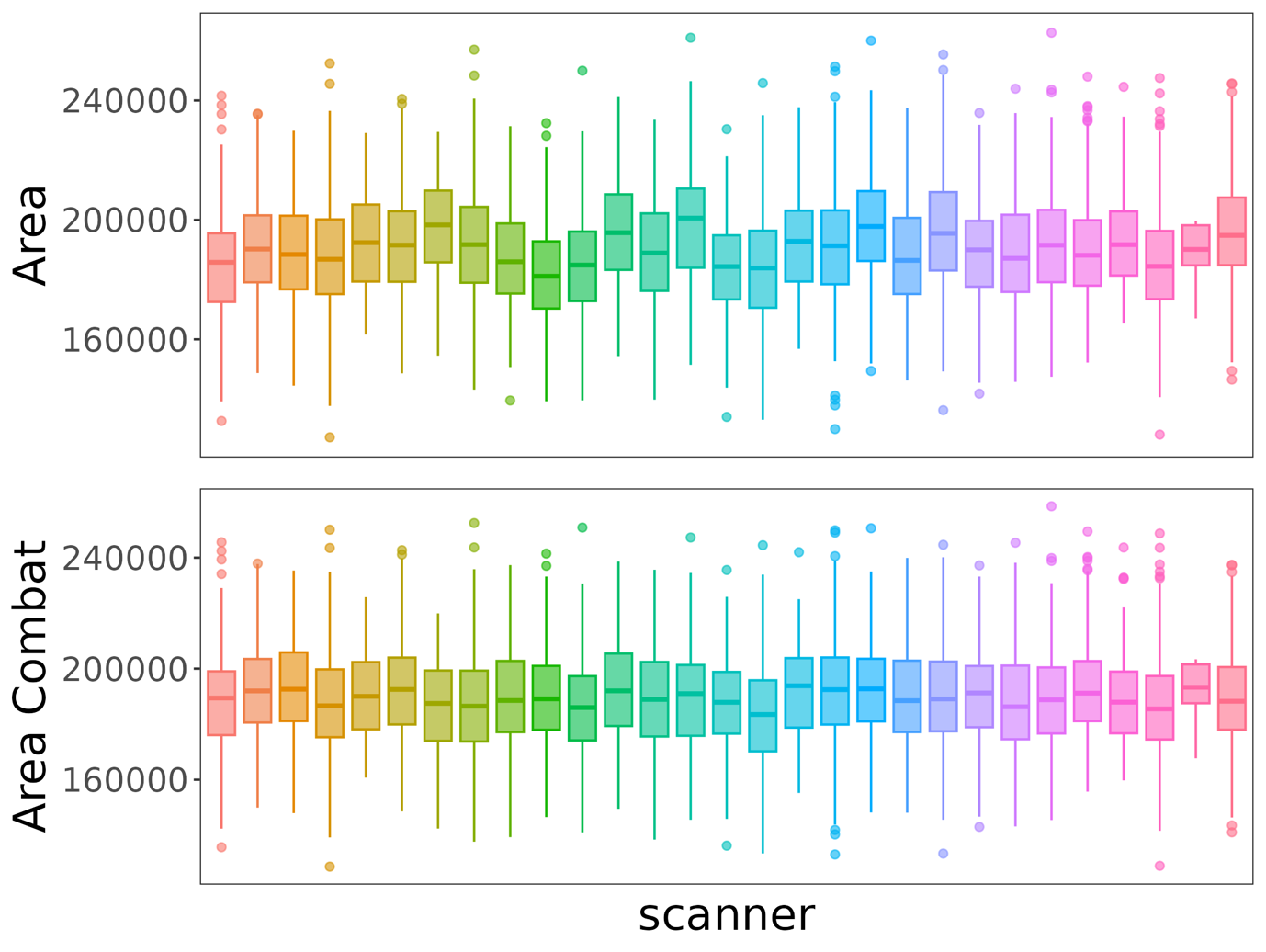
 SI Figure 8. Box plots of total surface area across scanners pre- and post harmonization. The figure shows box plots of the distribution of total surface area across scanners pre-scanner harmonization at the top, and post-harmonization at the bottom

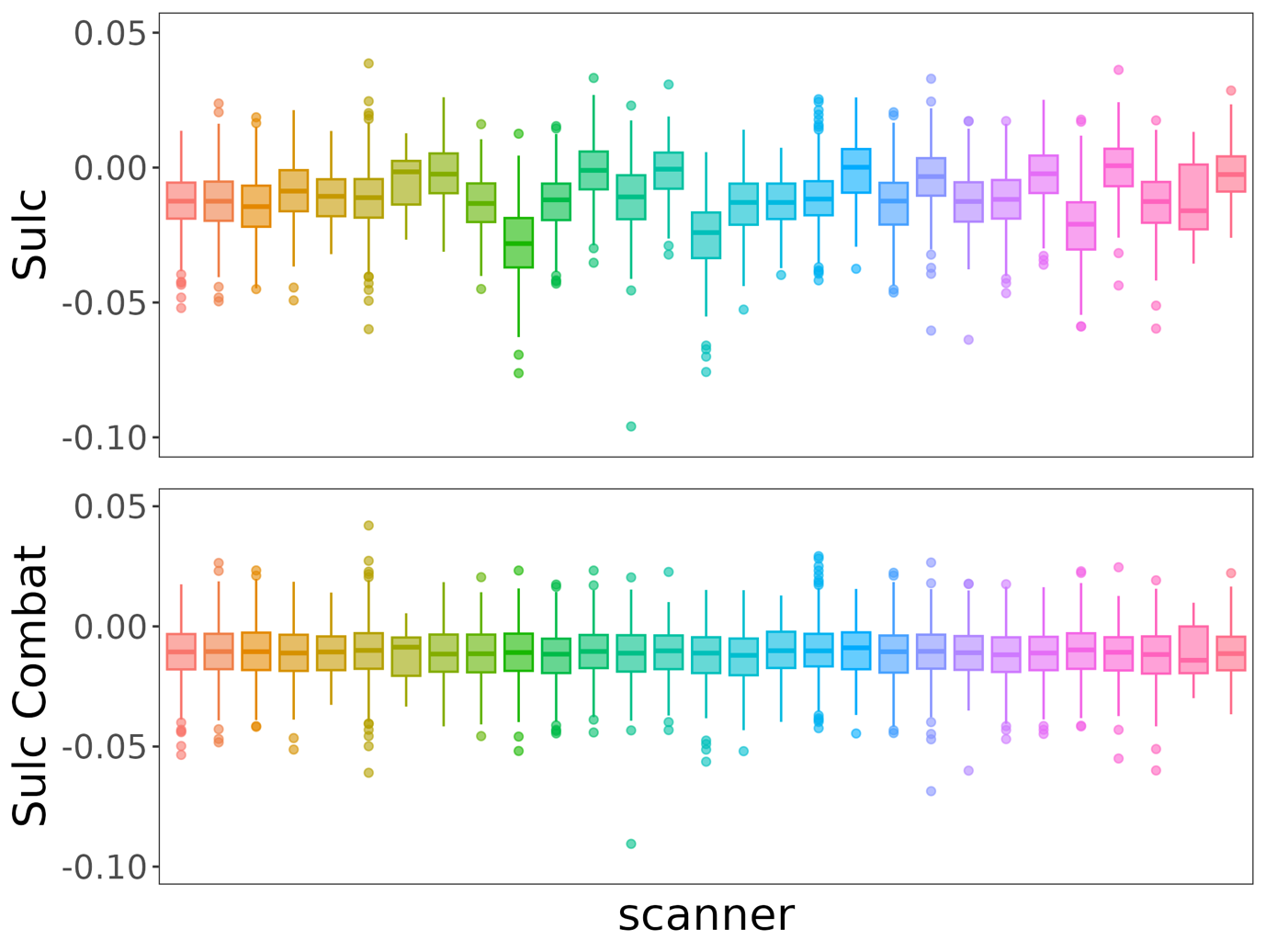

SI Figure 9. Box plots of mean sulcal depth across scanners pre- and post harmonization. The figure shows box plots of the distribution of mean sulcal depth across scanners pre-scanner harmonization at the top, and post-harmonization at the bottom.

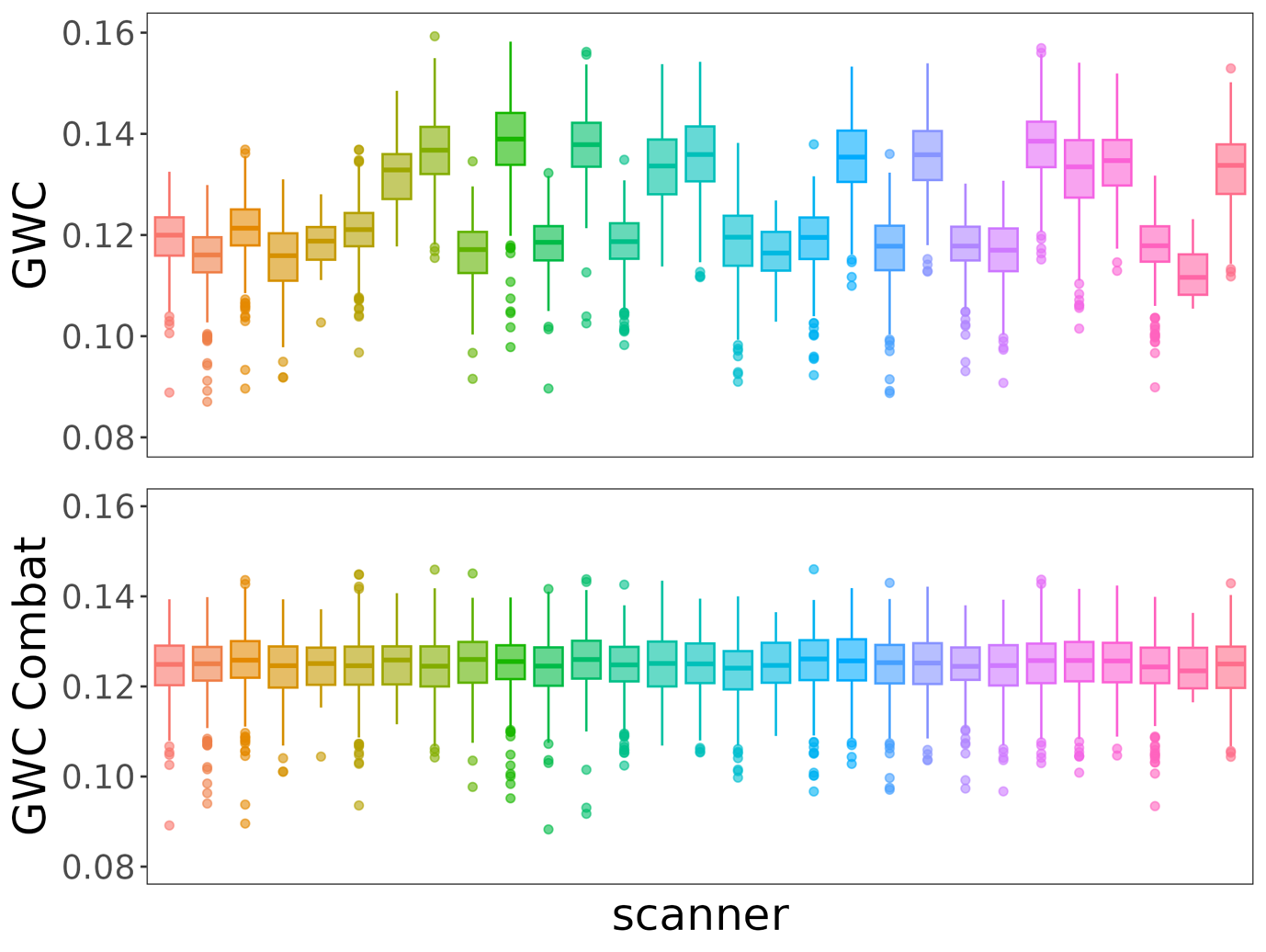

SI Figure 10. Box plots of mean grey/white-matter contrast (GWC) across scanners pre- and post harmonization. The figure shows box plots of the distribution of mean GWC across scanners pre-scanner harmonization at the top, and post-harmonization at the bottom.
